# Positive end-expiratory pressure versus sham valve/zero end-expiratory pressure in cardiopulmonary resuscitation during manual ventilation to improve neurological outcomes in adult patients suffering an out-of-hospital cardiac arrest – an investigator-initiated, pragmatic, registry-based, multicenter, parallel-group, triple-blind randomized controlled superiority clinical trial in the ARREST registry (REVIVE-PEEP protocol Stage-1 Registered Report)

**DOI:** 10.64898/2026.08.27.26361533

**Authors:** Jeroen van Eijk, Patrick Schober, Hans van Schuppen, Judith ter Schure

**Affiliations:** Department of Anesthesiology, Amsterdam UMC, Amsterdam, The Netherlands; Amsterdam Public Health, Amsterdam, The Netherlands; Department of Epidemiology & Data Science, Amsterdam UMC, Amsterdam, The Netherlands

**Keywords:** positive end-expiratory pressure (PEEP), zero end-expiratory pressure (ZEEP), cardiopulmonary resuscitation (CPR), out-of-hospital cardiac arrest (OHCA), oxygenation, ventilation, emergency medical service (EMS), ARREST registry

## Abstract

**Background:** In patients experiencing out-of-hospital cardiac arrest, optimizing oxygen delivery to vital organs during cardiopulmonary resuscitation (CPR) is critical. Ventilation is an essential component of CPR, but the effects of different ventilation strategies on patient outcomes have not been adequately established. Both positive end-expiratory pressure (PEEP) and zero end-expiratory pressure (ZEEP) are used during CPR, but their comparative effectiveness remains unclear.

**Methods:** This investigator-initiated, pragmatic, registry-based, multicenter, triple-blind randomized controlled superiority trial evaluates whether applying 8 cm H_2_O PEEP during cardiopulmonary resuscitation (CPR) improves outcomes compared with ZEEP in adults with non-traumatic, non-drowning out-of-hospital cardiac arrest. Pre-randomized CPR kits (1:1 PEEP vs. sham) were used by ambulance sites during manual ventilation throughout the resuscitation process. The main analyses were conducted in the modified intention-to-treat (mITT) subset of patients who received no sustained mechanical ventilation and either a supraglottic airway or endotracheal tube.

The primary outcome was neurological status at hospital discharge measured by a utility-weighted score on the modified Rankin Scale, analyzed in a regression model with main effects for witnessed arrest, bystander CPR and shockable initial rhythm. Six prespecified interim analyses were performed on the primary outcome by a data safety monitoring board for both benefit and harm, on confidence intervals corrected for sequential analysis. Secondary outcomes included any return of spontaneous circulation during the CPR attempt, 30-day survival, and 6-month quality of life. The primary safety outcome was clinically significant pneumothorax. Data were collected within the existing ARREST (AmsteRdam RESustitation STudies) registry.

**Results:** Among the 3,200 patients who underwent randomization, 1,200 in the PEEP group and 1,200 in the ZEEP group were included in the mITT analysis. The mean difference in utility-weighted scores on the modified Rankin Scale, corrected for covariates, was 0.0073 ; 95% CI -0.0134–0.0279, corrected for prespecified interim analyses. The uncorrected mean utility was 0.1020 in the PEEP group and 0.1075 in the ZEEP group. The observed rates in the PEEP and ZEEP groups of any return of spontaneous circulation were X% vs. Y%; adjusted odds ratio, Z; 95% CI, A–B and in 30-day survival X% vs. Y%; adjusted odds ratio, Z; 95% CI, A–B. The incidence of clinically significant pneumothorax was similar between the PEEP and ZEEP groups, occurring in X% and Y% of patients, respectively (adjusted odds ratio, Z; 95% CI, A–B).

**Conclusion:** Conclusion and discussion follow at Stage-2.

**Stage-1 Registered Report:** We present our Stage-1 Registered Report as a full clinical trial article with all methods in past tense. Mock results, table and figures are included for the primary analysis, to provide full clarity about our intended analyses. To remind the reader that this Stage-1 article is written before data collection, we highlight in color that these mock results are only for illustrative purposes and will be replaced by the actual results in the Stage-2 Registered Report.

*Supplements:* All study supplements and updates are available in the replication package ResearchEquals collection [1], including the documents that received ethics approval – protocol Version 2.4 (September, 2026) [2], Patient Information Sheet [3], DSMB charter [4] and an example DSMB closed session report [5], as well as an Excel sheet [6] and R code [7] used to generate Figure A.5.

*Simultaneous evaluation PCI-RR and ethics:* The institutional review board (METC NedMec+)^1^ tasked with the ethical evaluation of the REVIVE-PEEP trial agreed to perform a pilot experiment of simultaneous ethical and Stage-1 peer review at Peer Community In Registered Reports (PCI-RR). This means that all feedback was considered as one round of review, the author’s reply to both weighted the two sources of feedback together and all review as made available on PCI-RR. All openly available supplements in the replication package [1] were also referenced in the grant review process for Clinical Study Program funding by the Dutch Heart Foundation ^2^, but grant review was not part of the pilot experiment. Nevertheless, we will evaluate the process as a major step towards ‘Trinity review’ aiming to reduce ‘redundant paperwork of three different documents – research papers, ethics review applications, and research grant applications – for the same research plan’[8].

*Trial registration:* The REVIVE-PEEP trial was registered (July 27, 2026) in the Netherlands registry OMON (‘Overzicht van Medisch-wetenschappelijk Onderzoek in Nederland’) under identification number NL-OMON61400^3^, with OMON providing the data to the International Clinical Trial Registry Platform.

*Protocol and statistical analysis plan:* This Stage-1 Registered Report (RR) serves as the full pre-registration alongside the protocol Version 2.4 (September, 2026) [2]. To illustrate the unique possibilities of a Registered Report for a clinical trial positioned within an ongoing registry, and to prepare all reporting decisions in detail for very fast completion of the eventual result publication, we intend to supplement this manuscript in the future with a full synthetic mock dataset. This mock data will be generated based on ARREST registry data (2016–2018) [9] and prepare code for all analyses, also for those that need less detail than the preregistered primary analysis. The mock data project will drive further details of a possible statistical analysis plan, but should not deviate from this Stage-1 RR manuscript. If anything does change, we will contact PCI-RR. If changes are minor, we will transparently flag the deviations in the Stage-2 manuscript. If the changes are major and require re-review, than the in-principle acceptance will be rescinded, possibly reinviting the reviewers to evaluate the changes, and a new in-principle acceptance issued in the event of approval. Although we will be still blinded to the allocation, the first data will have been collected in this case, which might affect the level of bias control.

*Funding:* The legal sponsor of the trial was Amsterdam UMC, and the trial was coordinated by the ARREST registry. The Dutch Heart Foundation awarded funding for the trial through its Clinical Study Program (01-001-2026-0740). Study products were provided by Ambu A/S (Ballerup, Denmark), as in-kind support and technical expertise for the development, production, and regulatory preparation of the blinded study devices. Stryker (Redmond, WA, USA) supports the broader ARREST registry infrastructure through a separate grant. Neither company was involved in the scientific conduct, analysis, interpretation, or publication of the trial.

## 1. Introduction

Out-of-hospital cardiac arrest remains a major global health burden, with approximately four million cases annually, including around 17,000 in the Netherlands. Although 30-day survival rates have improved modestly to approximately 20% today in the Netherlands, these rates have plateaued in the past decade [10]. This stagnation underscores the urgent need for evidence-based interventions to further improve advanced life support practice.

The fundamental goal of cardiopulmonary resuscitation (CPR) is to ensure adequate supply of oxygenated blood to the brain and heart until spontaneous circulation is restored [11]. Although chest compressions are essential to maintain cardiac output, effective ventilation is equally critical to maintain oxygenation. Hypoxemia during CPR is common, and observational studies have shown an association with reduced probability of successful resuscitation and poorer neurological outcomes [12, 13, 14].

Despite international guidelines emphasizing the importance of effective ventilation during resuscitation, there is no high-quality evidence to support specific ventilation parameters during cardiac arrest [15]. A systematic review by the International Liaison Committee on Resuscitation identified the lack of randomized controlled trials evaluating the effect of ventilation strategies on neurological outcomes as a key knowledge gap [16].

Positive end-expiratory pressure (PEEP) plays a fundamental role in optimizing oxygenation in routine critical care practice, but its role during cardiac arrest remains largely unexplored. To date, no clinical trial has evaluated its effect in this context, although two pilot studies illustrate growing clinical interest (*see Appendix Section A.1.3*). Conversely, negative intrathoracic pressure during conventional CPR has not been shown to improve neurological outcome. [17]. As PEEP represents its physiological opposite, its effects during CPR warrants investigation.

Animal studies have demonstrated the biological plausibility of PEEP in improving oxygenation during CPR [18, 19, 20]. Despite its potential importance, there is currently no consensus on the use of PEEP in clinical practice, resulting in significant variation in its application during cardiac arrest [21].

To address this, we designed the Resuscitation and Ventilation Innovations Via Evidence-based Positive End-Expiratory Pressure (REVIVE-PEEP) trial. The objective of this study was to evaluate whether the application of 8 cm H_2_O of PEEP, compared to a sham valve applying zero end-expiratory pressure (ZEEP), during manual ventilation after advanced airway management in out-of-hospital cardiac arrest improves neurological outcomes at discharge.

## 2. Methods

### 2.1 Trial design and oversight

The REVIVE-PEEP trial, an investigator-initiated, pragmatic, registry-based, multicenter, parallel-group, triple-blind 1:1 individually randomized controlled superiority clinical trial was conducted from October 2026 to March 2029 within the ARREST (AmsteRdam RESustitation STudies) registry. The ARREST registry is a longstanding cardiac arrest registry currently encompassing five emergency medical services in the Netherlands, serving a population of approximately 3.5 million people [9]. The term pragmatic in REVIVE-PEEP refers to the extent the trial evaluates the intervention under usual-care conditions and should not be interpreted as implying reduced methodological rigor or data quality. Specifically, the pragmatic nature of REVIVE-PEEP is most strongly reflected in the domains of eligibility, recruitment, setting, organization, flexibility of intervention delivery, and the primary outcome, whereas follow-up and the primary analysis are less pragmatic. The trial characteristics across the pragmatic–explanatory continuum are specified using the PRECIS-2 framework [22] in Appendix Section A.1.1.

The trial protocol was previously published, and the first draft of this manuscript was published as a registered report [23] to enhance transparency and methodological rigor. The Medical Research Ethics Committee NedMec+ reviewed and approved the study in accordance with the European Medical Device Regulation (MDR; EU 2017/745), the Dutch Medical Research Involving Human Subjects Act (WMO), and other applicable legislation (July 22, 2026; NL-009273).

Due to the emergency nature of the study, a deferred informed consent was approved. Participants were informed about their participation during hospital admission, preferably once they had regained sufficient decision-making capacity, and written informed consent was obtained from the participant or, if they remained incapacitated, their legal representative. If consent could not be obtained during hospital admission, participants or their legal representatives were contacted directly after discharge.

A steering group, including patient representatives, designed the trial and developed the protocol (available as a supplement [2]). Data analyses were performed by Jeroen van Eijk and Judith ter Schure, and verified by Patrick Schober, all vouching for the accuracy and completeness of the data and for the fidelity of the trial to the protocol. An independent Data and Safety Monitoring Board (DSMB) oversaw the trial, reviewing data at six prespecified interim analyses. The DSMB monitored safety and applied prespecified stopping criteria for efficacy, including both benefit and harm (details in [4] and [5]).

#### Changes to the protocol

None.

### 2.2 Trial setting

In the Netherlands, two ambulances respond to suspected OHCA cases, both staffed by a driver and an ALS ambulance nurse, trained in manual defibrillation, advanced airway management, drug administration, and vascular access. Ambulance nurses provide prehospital care according to Dutch national guidelines, based on the European Resuscitation Council guidelines.[15] The ambulance nurses are allowed to terminate resuscitation on scene, based on established criteria.

### 2.3 Patient population eligibility criteria

Patients 18 years of age or older were eligible for inclusion in the trial after advanced airway management with an endotracheal tube or a supraglottic airway device (i-gel®, Intersurgical, Wokingham, UK) during an out-of-hospital cardiac arrest. Exclusion criteria were a suspected traumatic etiology of the cardiac arrest, suspected drowning or the use mechanical ventilation. See Figure 1 for an overview of randomization and patient inclusion.

**Figure 1.**
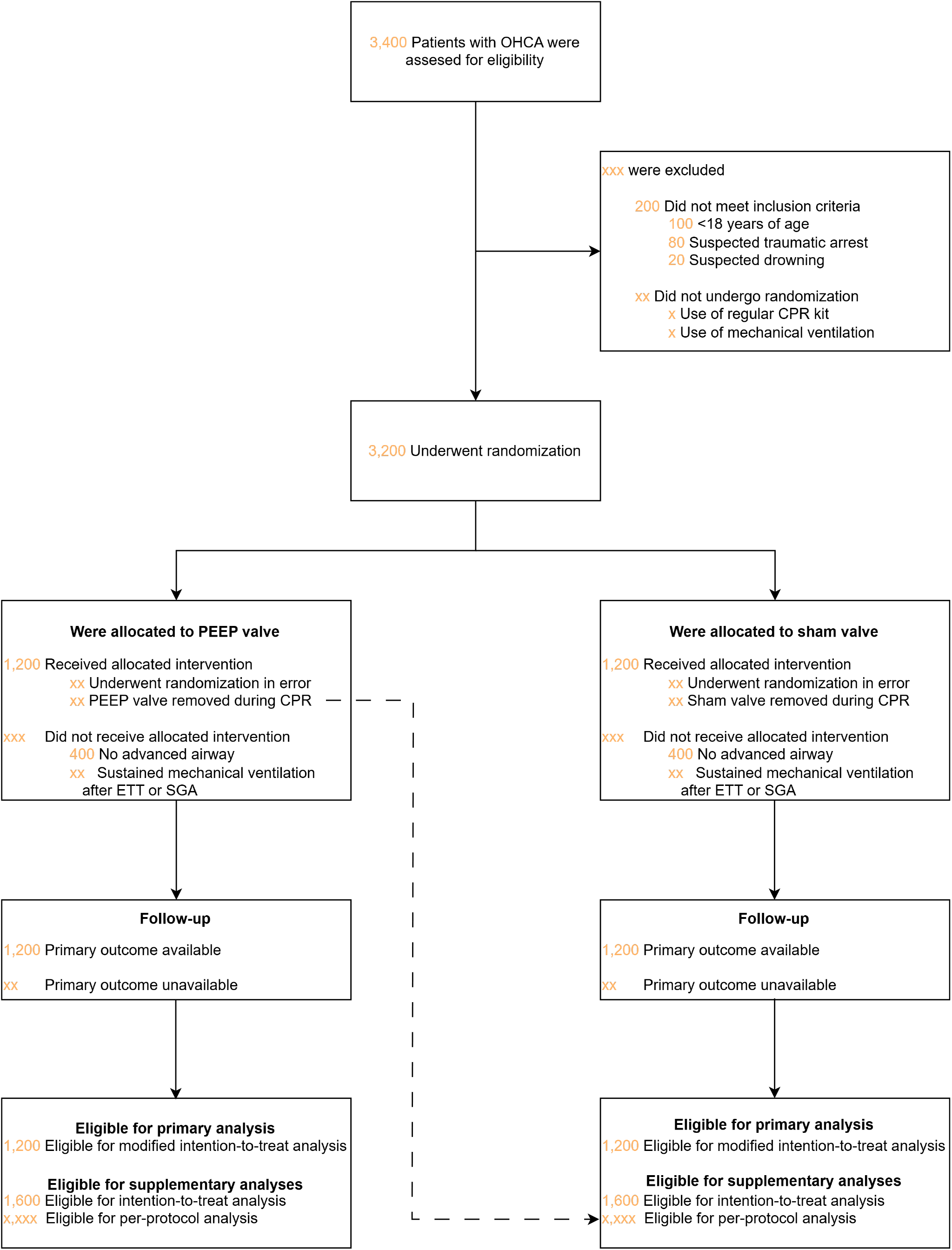
Design flow diagram with mock sample sizes. The modified intention-to-treat analysis subset is defined as those patients receiving CPR with the allocated study bag, whether or not the valve is removed, with manual ventilation (no sustained mechanical ventilation) after advanced airway management, i.e. endotracheal tube (ETT) or supraglottic airway device (SGA).

### 2.4 Intervention and comparator

During the study, ventilation was performed using a standard disposable resuscitator bag (SPUR II, Ambu A/S, Ballerup, Denmark) with a pre-assembled blinded PEEP or sham (ZEEP) valve, provided as a single resuscitator kit. Patients were considered randomized once ventilation with the study device commenced during CPR. The resuscitator bag from the first-arriving ambulance was used throughout the resuscitation attempt, both for bag-valve-mask ventilation and after advanced airway management.

The intervention valve provided 8 cm H_2_O of PEEP during manual ventilation, while the sham valve in the control group mimicked the intervention valve in appearance, but remained inactive, delivering ZEEP. Both valves were non-adjustable and identical in size, shape, and external markings (*see Appendix A.1.2 Dose and investigational product*).

Ambulance personnel were familiar with the use of PEEP valves as part of standard care. They received an informational video explaining the study and completed a mandatory e-learning. If necessary, they could consult local study ambassadors with questions. In the case of sustained ROSC (following definitions from [25]), ambulance personnel may remove the PEEP valve at their discretion if deemed clinically necessary. For patient categories excluded from the study, such as drowning or suspected trauma cardiac arrest, the study valve can be removed and possibly replaced by a standard adjustable non-blinded CE-marked PEEP valve. For any other reasons, such as the clinical impression of elevated ventilation pressures or thoracic hyperinflation, for example in patients with severe asthma, the removal of the valve was at the discretion of the ambulance personal and noted in the case report form.

### 2.5 Outcomes and harms

The primary outcome of the study was the neurological outcome at hospital discharge, assessed using a utility-weighted modified Rankin Scale that takes into account patient preferences and the functional impact of recovery on quality of life (Table 1) [24].

**Table 1.** Utility weights for the modified Rankin Scale from Chaisinanunkul et al., 2015 [24].

| Score | Utility weight | Definition |
| --- | --- | --- |
| 0 | 1 | **No symptoms** |
| 1 | 0.91 | **No significant disability**<br>Able to carry out all usual activities, despite some symptoms |
| 2 | 0.76 | **Slight disability**<br>Able to look after own affairs without assistance, but unable to carry out all previous activities |
| 3 | 0.65 | **Moderate disability**<br>Requires some help, but able to walk unassisted |
| 4 | 0.33 | **Moderately severe disability**<br>Unable to attend to own bodily needs without assistance or unable to walk unassisted |
| 5 | 0 | **Severe disability**<br>Requires constant nursing care and attention, bedridden, incontinent |
| 6 | 0 | **Death** |

Key secondary outcomes included any return of spontaneous circulation (ROSC) during the CPR attempt, 30-day survival, favorable neurological outcome (defined as a score of 0 to 3 on the modified Rankin Scale), and health-related quality of life at 6 months among survivors, as assessed by the EuroQol Group 5-Dimension 5-Level questionnaire. All outcome definitions are in agreement with 2024 update of the Utstein out-of-hospital cardiac arrest registry template [25].

The primary safety outcome is the occurrence of clinically significant pneumothorax, defined as the need for drainage via (needle) thoracostomy or chest tube. In a subset of patients, advanced respiratory parameters were collected to evaluate the effect of the intervention on pulmonary mechanics. This includes assessing peak pressures, effective PEEP, and lung compliance.

### 2.6 Sample size

We defined an absolute risk difference of 4% in mortality as minimally clinically important (*see PCI-RR design planner table Section A.1.9*) and considered two scenarios for the neurological outcomes of these additional 4% survivors, a Favorable scenario and an Unfavorable scenario (*see Appendix Section A.1.7 and Figure A.5*).

A sample size of 2,400 patients was specified based on a two-sided test at level alpha of 5% and a range of Cohen’s d 0.117-0.137 corresponding to a minimum under the Unfavorable scenario and a maximum in the Favorable scenario. With six interim analyses performed by the DSMB [4] at 1,000; 1,200; 1,400; 1,600; 1,800; and 2,000 patients following an O’Brien-Fleming alpha-spending function, this trial aimed for 81%-91% power (see Appendix Figure A.4).

Because the primary analysis was restricted to the mITT subset, and approximately 25% of randomized patients were expected to have ROSC before advanced airway management or to receive sustained mechanical ventilation, 3,200 patients were randomized to include the required 2,400 patients in the primary analysis.

### 2.7 Randomization and blinding

The randomization list was generated using Castor Electronic Data Capture system (Castor v2026.1.5.0 Castor CDMS, data.castoredc.com), with unstratified randomly permuted blocks of randomly chosen size two and four in a 1:1 ratio. This procedure was known within the study team, openly available online in this document and the protocol [2]) but was not communicated to the ambulance personnel in an instruction manual or e-learning. Study products were assembled and labeled according to the list, repacked in sealed, identically labeled bags in batches of 12, and distributed to ambulance posts. Restocking ensured patient-level randomization unknown to the ambulance personnel that enrolled participants by using the prerandomized kits. Unblinding was permitted only when clinically and statistically indicated by the DSMB. Access to the randomization list was restricted to the research data management department that exported the randomization list, the medical technical support staff responsible for packing and distribution, and the DSMB statistician that also served as the independent statistician to generate the DSMB closed session reports. Access to the original list in Castor, in case of problems that require verification, was given to two users profiles from the research data management department, one user profile from the medical technical department, and – only if settings would be changed to unblinded (visible in the audit trail) – to the user profile of the trial statistician Judith ter Schure. The trial was triple-blind, and the allocation of treatment was concealed from ambulance personnel, participants, outcome assessors, and trial steering group (including the trial statistician) throughout the study.

### 2.8 Statistical analysis

In the primary analysis, a linear regression model was used to estimate the mean difference in utility-weighted modified Rankin Scale, adjusted for important prognostic factors, i.e. whether the cardiac arrest had been witnessed, whether cardiopulmonary resuscitation had been initiated by a by-stander, and an initial shockable rhythm (*see Appendix Section A.1.4*). Results are reported as 95% confidence intervals for the mean difference in utility, based on a Gaussian approximation and the O’Brien-Fleming alpha-spending function (*see Appendix Figure A.4*). All statistical analyses were performed in R and analysis code is available in the replication package [1].

#### Intercurrent events and estimands

The primary analysis was performed on the modified intention-to-treat (mITT) subset defined by three different intercurrent events that are illustrated in Figure 1: (a) removal of the study valve during CPR, (b) no initiation of advanced airway management and (c) sustained mechanical ventilation after advanced airway management. The mITT subset includes the patients that had their randomized intervention discontinued by removing the valve (a) – targeting a treatment policy estimand – and excludes patients that did not receive the randomized intervention due to no advanced airway or sustained mechanical ventilation (b and c) – targeting a principal stratum estimand (*see Appendix Section A.1.6*).

#### Sensitivity and supplementary analyses

As a sensitivity analysis for the primary outcome in the mITT subset, we estimate the mean difference in utility-weighted modified Rankin Scale that is not corrected for covariates. The intention-to-treat (ITT), per-protocol (PP) analyses and the analysis on the three non-mITT subsets (no advanced airway due to ROSC, no advanced airway due to stopped CPR, and sustained mechanical ventilation) are reported as supplementary analyses (Figure 1; *see Appendix Section A.1.6*).

### 2.9 Prespecified possible conclusions

We prespecified three possible primary conclusions:

I. The confidence interval for the mean difference in utility includes only positive values that indicate a benefit of PEEP. In this case, we will recommend to implement 8 cm H_2_O PEEP in CPR in adult patients suffering an OHCA not related to trauma or drowning, also in case of a small effect – given that cost and burden of implementation are low, while gain of each percentage point benefit is high, equaling hundreds of lives saved in The Netherlands and ten thousands worldwide.
II. Either the confidence interval for the mean difference in utility includes only negative values that indicate harm of PEEP, or we find safety concerns on the safety outcome of clinically significant pneumothorax. In this case, we would recommend against routine implementation of manual PEEP at 8 cm H_2_O during CPR. These findings would not preclude further investigation of alternative PEEP levels in future studies.
III. We find no safety concerns and an inconclusive result with effects of benefit and harm both in the confidence interval. In this case, we recommend further studies to draw conclusions in a meta-analysis, while taking into account feasibility (e.g. value of information and clinical relevance of the values within the confidence interval).

## 3. Results

### Study participants

Of the 3,400 patients screened for eligibility, 3,200 were randomized, with the reasons for exclusion outlined in Figure 1.

Among the randomized patients, 2,400 were included in the mITT subset. Of these, 1,200 patients received PEEP and 1,200 patients received ZEEP. The intention-to-treat subset also includes an additional [X] patients who had ROSC prior to advanced airway, [Y] patients who had their CPR attempt stopped before advanced airway management and [Z] who had sustained mechanical ventilation during resuscitation, see Figure 1.

The patient characteristics of both groups, including the subset ineligible for mITT analysis, are reported in *Table not included*.

### Outcomes

#### Primary outcome

Data for the primary outcome were available for 1,200 patients (100%) in the PEEP group and 1,200 patients (100%) in the ZEEP group. A total of 156 patients in the PEEP group and 162 patients in the ZEEP group survived to hospital discharge, as shown in Figure 2. Their neurological outcomes, as assessed using the utility-weighted modified Rankin Scale, are presented in Table 2. The primary analysis was inconclusive regarding the effect of PEEP on neurological outcomes, with a mean difference of 0.0073 (95% CI corrected for sequential analysis: -0.0134–0.0279). Figure 3 shows the full sequential analysis over the six interim analyses and final analysis in comparison to the two scenarios used in the power analysis. Sensitivity analyses conducted on the same mITT subset uncorrected for covariates demonstrated consistent findings. *No mock results are provided for sensitivity and supplementary analyses but details on the primary analysis are provided in the Appendix Section A.2*.

**Table 2.** Neurological outcomes on the modified Rankin Scale weighted by Chaisinanunkul et al. (2025) [24] utility weights: difference in mean utility (not corrected in a regression model) at the final analysis of 2,400 patients in the mITT subset with mock results illustrated by two routes of calculations. Top: first weighting absolute proportion differences and then taking the combined sum; Bottom: first weighted average (mean utility) and then taking the difference.

| mRS score | PEEP | ZEEP/sham | Absolute proportion diff | Utility weight | Weighted diff |
| --- | --- | --- | --- | --- | --- |
| 0 | 19 (1.58%) | 25 (2.08%) | -0.0050 | 1 | -0.0050 |
| 1 | 65 (5.42%) | 63 (5.25%) | 0.0017 | 0.91 | 0.0015 |
| 2 | 33 (2.75%) | 46 (3.83%) | -0.0108 | 0.76 | -0.0082 |
| 3 | 26 (2.17%) | 16 (1.33%) | 0.0083 | 0.65 | 0.0054 |
| 4 | 7 (0.58%) | 4 (0.33%) | 0.0025 | 0.33 | 0.0008 |
| 5 | 6 (0.50%) | 8 (0.67%) | -0.0017 | 0 | 0 |
| 6 | 1,044 (87.0%) | 1,038 (86.5%) | 0.0050 | 0 | 0 |
| Combined | 1,200 (100.%) | 1,200 (100.%) |  |  | -0.0055 |
| Mean utility (SD) | 0.1020 (0.2757) | 0.1075 (0.2845) |  |  | -0.0055 |
| (Std. Error) | (0.0080) | (0.0082) |  |  | (0.0114) |

**Figure 2.**
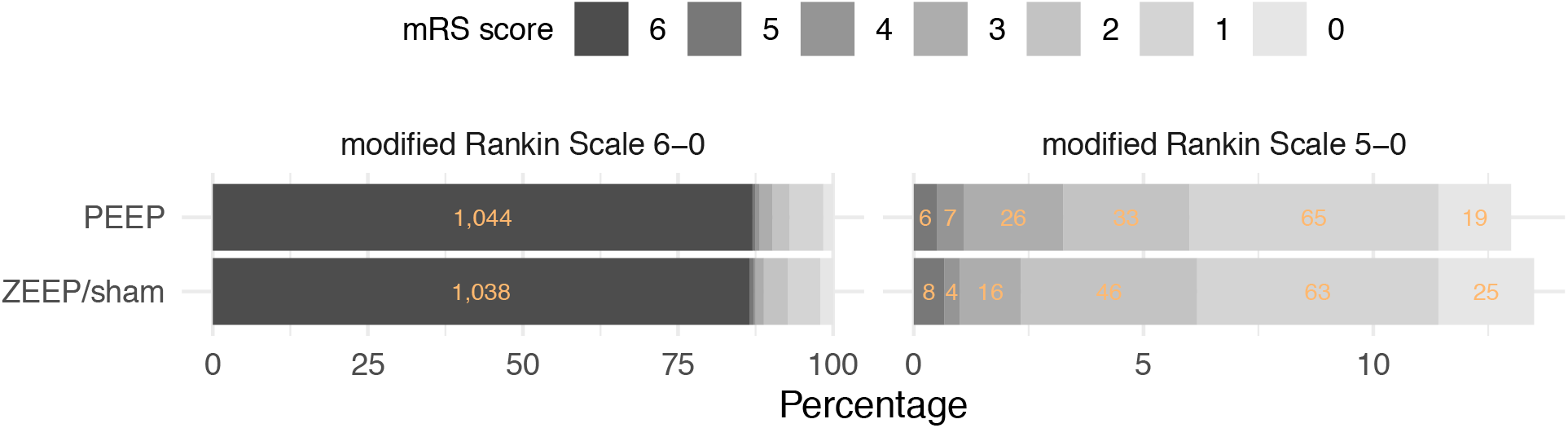
Neurological outcomes on the modified Rankin Scale at the final analysis of 2,400 patients with mock results. 6: Death – 5: Severe disability – 4: Moderately severe disability – 3: Moderate disability – 2: Slight disability – 1: No significant disability – 0: No symptoms. Left: All patients. Right: Zoom on the patients that survived (excluding mRS-6).

**Figure 3.**
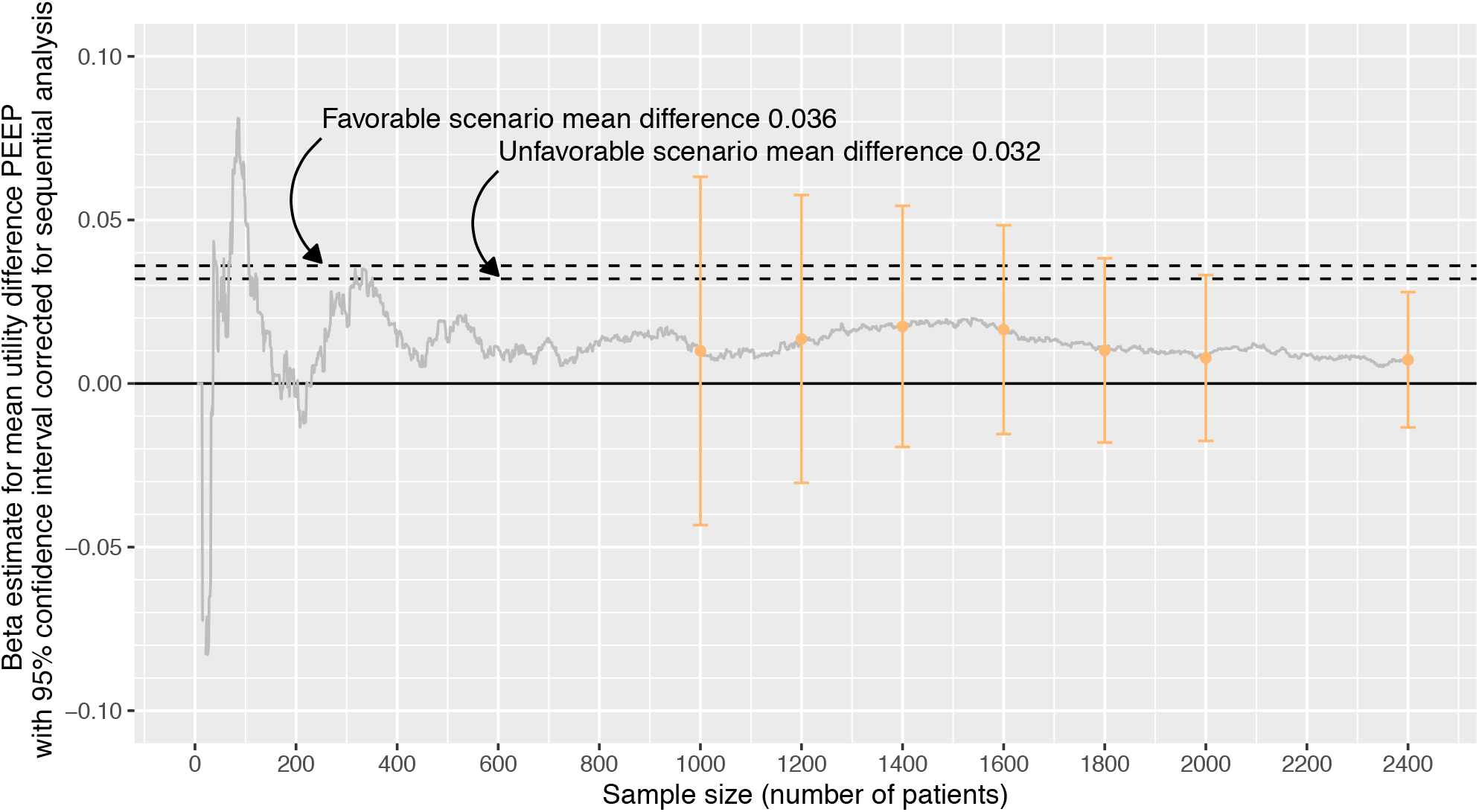
Neurological outcomes on the modified Rankin Scale weighted by Table 1: interim analyses over time with mock results. *See Figure A.5 for an illustration of the mean differences from the Favorable and Unfavorable scenario*.

### Secondary outcomes

There was no evidence of a significant difference between the PEEP group and the ZEEP group in the proportion of patients who achieved return of spontaneous circulation. In the PEEP group, [number PEEP group] patients ([percentage PEEP group]%) achieved any return of spontaneous circulation during the CPR attempt, compared to [number ZEEP group] patients ([percentage ZEEP group]%) in the ZEEP group (adjusted odds ratio [aOR], [value]; 95% CI, [lower bound] to [upper bound]). Similarly, survival at 30 days did not differ significantly between the two groups. Survival was observed in [number PEEP group 30d] patients ([percentage PEEP group 30d]%) in the PEEP group and [number ZEEP group 30d] patients ([percentage ZEEP group 30d]%) in the ZEEP group (aOR, [value]; 95% CI, [lower bound] to [upper bound]). Additionally, the proportion of patients with favorable neurological outcomes, as dichotomized on the modified Rankin Scale, was similar between the PEEP and ZEEP groups. In the PEEP group, [number PEEP MRS] patients ([percentage PEEP MRS]%) had favorable neurological outcomes, compared to [number ZEEP MRS] patients ([percentage ZEEP MRS]%) in the ZEEP group (aOR, [value]; 95% CI, [lower bound] to [upper bound]). There was also no significant difference between the two groups in quality of life at 6 months, with mean scores of [mean PEEP group] and [mean ZEEP group] (aOR, [value]; 95% CI, [lower bound] to [upper bound]).

### Subgroup analyses

No mock results on subgroup analyses provided in this Stage-1 Registered Report. Actual results will follow in Stage-2.

### Physiological parameters

No mock results on physiological parameters provided in this Stage-1 Registered Report. Actual results will follow in Stage-2.

## 4. Discussion

The observations on our safety outcome of clinically relevant pneumothorax are quite possibly incomplete, as the clinical examination including auscultation is known to have limited sensitivity [26, 27]). However, the lack of difference between the PEEP and ZEEP/sham group observed in this trial is in agreement with the results on the primary outcome of neurological outcomes on the modified Rankin Score. *Further discussion will follow actual results in Stage-2*.

## Data Availability

All data produced will be made available in the Replication package (for reproducibility and replication) at ResearchEquals: https://doi.org/10.53962/p8t5-wzwr

## Acknowledgments

We thank the patient representatives, ambulance and hospital personnel, EMS medical directors and board members of the participating emergency medical services, DSMB members and our co-authors on the REVIVE-PEEP trial protocol and other supplements, Michiel Hulleman, Stephan Loer, Lothar Schwarte, Thijs Delnoij, Mette Ekkel and Remy Stieglis, for their dedication to the trial. We also thank the institutional review board, METC Amsterdam UMC, for supporting the pilot of simultaneous review of ethics and Stage-1 Registered Report, and the Dutch Heart Foundation for funding. We are grateful to PCI-RR founder and managing board member Chris Chambers for his detailed and thoughtful responses to our questions about the PCI-RR platform, and Anna Porter (Recommender), Nicolas Segond (Reviewer) and David Purkarthofer (Reviewer) for the smooth and insightful review process. We acknowledge the PCI-RR snapshot template for stressing the importance of a-priori specifying the conclusions that will be drawn given different results (Section 2.9), Berry Consultants for putting us on the right track of utility weights for the modified Rankin Scale with their “In the Interim… “ podcast episodes 5, 37 and 50 [28, 29, 30], and Yongxi Long and Erik van Zwet for their advice and expertise [31] on the analysis of neurological outcomes. We credit Caroline for the design of the REVIVE-PEEP logo that appears alongside the review at PCI-RR.

Generative AI (Large Language Models) was used to accelerate R coding for figures, and to improve writing at the level of short paragraphs.

## Data and software availability

An Excel sheet is provided as supplementary material [6] that calculates Cohen’s d from the scenarios presented in Appendix Figure A.5 and allows to try custom new scenarios. R code is provided [7] that takes the input from the Excel sheet to reproduce Appendix Figure A.5 in R studio [32], using the package ggplot2 [33], dplyr [34] and gridExtra [35]. nQuery was used for the sample size calculation (see Figure A.4). We used the LateX template [36] by Mathias Legrand that was made available for Overleaf.

## 1. Appendix

### A.1 Methods details

#### A.1.1 Trial characteristics across the pragmatic-explanatory continuum

The REVIVE-PEEP trial is a pragmatic trial, based on the following domains scored according to PRECIS-2 [22]:

**Table A.3.** PRECIS-2 Assessment for REVIVE-PEEP Trial. Scores range from 1 (very explanatory) to 5 (very pragmatic).

| PRECIS-2 domain | Score | Rationale for REVIVE-PEEP |
| --- | --- | --- |
| Eligibility | 4 | Broad inclusion of adult patients with non-traumatic, non-drowning OHCA, without selection based on prognosis, initial rhythm, witness status, or bystander CPR. |
| Recruitment | 5 | Enrollment occurs during routine emergency care without a separate pre-enrollment recruitment procedure; consent is deferred because of the emergency setting. |
| Setting | 5 | The intervention is delivered in routine prehospital care by participating regional ambulance services. |
| Organisation | 4 | Usual EMS personnel and familiar ventilation equipment are used; additional requirements are limited mainly to pre-randomized study kits, logistics, and study-specific training. |
| Flexibility (delivery) | 4 | The randomized PEEP level is fixed, but otherwise ALS care, co-interventions, and clinical decisions remain according to usual protocols and clinical judgement. |
| Flexibility (adherence) | N/A | The intervention is delivered acutely by EMS professionals; participant adherence is therefore not applicable. |
| Follow-up | 3 | Most outcomes are obtained from the existing ARREST infrastructure and routine clinical records; additional structured follow-up is mainly required for the 6-month quality-of-life outcome and study-specific mechanistic/physiological outcomes. |
| Primary outcome | 5 | Neurological outcome at discharge measured using the utility-weighted mRS is a directly patient-relevant clinical outcome rather than a physiological or surrogate endpoint. |
| Primary analysis | 2 | The primary analysis is performed in a prespecified mITT subset targeting the principal-stratum estimand and therefore does not include all randomized participants; conventional ITT analysis is supplementary. |

#### A.1.2 Dose and investigational product

##### Literature and guidelines

Three animal experimental studies [18, 19, 20] show that PEEP levels ≤ 10 cm H_2_O have no significant influence on hemodynamics during resuscitation, while higher PEEP levels improve gas exchange. The latest ERC guideline recommends a PEEP of 0 to 5 cm H_2_O, but this is entirely based on expert opinion: the ILCOR working group [16] indicates that there are no empirical studies supporting this recommendation in patients suffering cardiac arrest out-of-hospital, although the use of 5 cm H_2_O is consistent with routine perioperative use in healthy patients.

##### Implications for trial Interpretation

Levenbrown et al. [18] do not present significant pairwise differences compared to the 10 cm H_2_O group in oxygen delivery, but suggest that 5 cm H _2_ O is optimal. This suggestion is based on exploratory clustering techniques and has not been confirmed by other studies. In our choice for 8 cm H_2_O instead of 5 cm H_2_O we weigh this signal against the informative value of the study at a higher dose: the primary aim of the trial is to convincingly demonstrate whether PEEP has an effect. We expect that of the three possible conclusions (benefit, harm and inconclusive, see Section 2.9), benefit and inconclusive are a-priori more likely than harm. Especially in an inconclusive study, a higher dose yields more valuable information, while a lower dose leaves the possibility that a new trial is needed at a higher dose.

##### Physiological considerations

Severe atelectasis occurs during CPR as a result of chest compressions, which makes a slightly higher PEEP than routinely used in healthy patients physiologically defensible to limit alveolar collapse. From a mechanistic perspective, it is essential that the PEEP is set above the lower inflection point of the pressure-volume curve. During prehospital resuscitations in patients, the median of the lower inflection point is 5.56 cm H_2_O (IQR 4.80–8.23 cm H_2_O) [37]. Hence, a PEEP of 5 cm H_2_O would be insufficient for approximately half of the patients, unlike the higher dose of 8 cm H_2_O.

##### Investigational PEEP valve

A PEEP level of 8 cm H_2_O is therefore the lowest value at which a clinically relevant effect can realistically be expected, without applying an unnecessarily high pressure. The PEEP valve used in this study has a tolerance of ±1.5 cm H_2_O due to passive resistance and pressure changes in the chest during compressions. With a set PEEP level of 5 cm H_2_O, the actual PEEP varies from 3.5–6.5 cm H_2_O; at 8 cm H_2_O, this ranges from 6.5–9.5 cm H_2_O.

##### Investigational sham valve

For the control group, an identically looking sham valve is used that is adapted to deliver 0 cm H_2_O of PEEP (i.e. ZEEP), with a tolerance of +0.3 cm H_2_O. This ensures the absence of clinically relevant PEEP while maintaining identical handling characteristics and appearance to the active intervention, thereby preserving blinding and minimizing performance bias.

#### A.1.3 Registered pilot trials

Two randomized controlled trials that study PEEP in OHCA are currently recruiting patients: the PerAVent trial in Germany [38, 39] and the Lazarus-PEEP trial in Belgium [40]. We consider both as pilot trials that illustrate the increasing interest in PEEP during CPR, but can neither provide the quality of evidence that REVIVE-PEEP is designed for, nor ask feasibility questions that need to be answered before starting REVIVE-PEEP.

##### Small sample size

The Lazarus-PEEP and PerAVent trial both have small sample size compared to the 3,200 patients we think need to be individually randomized to provide convincing evidence. Lazarus-PEEP reports a sample size of 132 patients, and PerAVent has a cluster-randomization approach that decreases the effective sample size of the 600 patients reported. These limitations are also acknowledged by the PerAVent authors writing that their trial is ‘designed primarily to assess the feasibility and preliminary effects of the intervention rather than to definitively establish superiority’ [38]. In contrast, the REVIVE-PEEP trial was designed to provide convincing evidence both in the scenarios of large effects and harm – allowing early stopping monitored by the DSMB, small effects – designed to randomize 3,200 patients with possible sample size re-estimation, and discouragement of future studies – with precise estimates that can convincingly inform the value of information of future trials in case clinical relevance of small effects is questionable.

##### No feasibility questions

REVIVE-PEEP will run within the ARREST registry and relies on extensive experience of research in this patient group as well as knowledge of data characteristics. Together with the ambulance sites’ familiarity with PEEP valves, these aspects of the REVIVE-PEEP trial leave no feasibility concerns that need to be addressed. A large patient-level randomized trial is specifically called for to research effects on neurological outcomes [16].

##### No joint project

We considered a joint project, especially given the prospective meta-analysis experience in our team, but currently decided against it. A joint analysis would be dominated by the REVIVE-PEEP data, such that the conclusion of the joint project would very likely be exactly the same as the one from REVIVE-PEEP alone. The benefits of the project would be small, while the costs in terms of effort for data harmonization (given differences in dose, primary outcome, blinding) and data sharing agreements would be considerable.

#### A.1.4 Main effect covariates for the primary analysis

Table A.4 details the definitions of the covariates prespecified as main effects in the linear model for the primary analysis and key secondary analyzes. (See Section A.2 for an example based on mock data.) These main effects are considered highly prognostic for favorable neurological outcomes and reduce background variation. Only a single main effect of PEEP is estimated, with no interaction terms in the regression (no separate PEEP effect estimate for different levels of the covariates). Such subgroup analyzes can be performed in Stage-2 and are considered exploratory.

#### A.1.5 Interim analyses

Results are reported as 95%-confidence intervals for the mean difference in utility based on a Gaussian approximation constructed at 1,000, 1,200, 1,400, 1,600, 1,800, 2,000 and 2,400 patients by margins of 3.280, 2.996, 2.766, 2.579, 2.427, 2.299 and 2.050 times the standard error respectively, shown as the upper efficacy bound in Appendix Figure A.4. In the Favorable scenario, the trial will already have 79% power at 2,000 patients and 13%, 27%, 43%, 58% and 70% power at the interim analyzes, giving a good chance of implementing a positive result a year earlier than the planned completion of the trial.

**Table A.4.** Definitions of the covariates prespecified as main effects in the linear model for the primary analysis.

| Covariate | Levels | Coding | ARREST code |
| --- | --- | --- | --- |
| Witness of CA | Unknown | Unknown/missing | COgetuiC 0 or N/A |
|  | No | No witness | COgetuiC 7 |
|  | Yes, EMS | Yes, by ambulance professional | COgetuiC 1 |
|  | Yes, non-EMS | Yes, by medical professional | COgetuiC 2 |
|  |  | Yes, by police department | COgetuiC 3 |
|  |  | Yes, by fire department | COgetuiC 4 |
|  |  | Yes, by partner | COgetuiC 5 |
|  |  | Yes, by family or friends | COgetuiC 6 |
|  |  | Yes, unknown by whom | COgetuiC 8 |
| Bystander CPR | Unknown | Unknown/missing | RAblsC 0 or N/A |
|  | No | No bystander CPR | RAblsC 6 |
|  | Yes | Yes, by partner | RAblsC 1 |
|  |  | Yes, by family member/friend | RAblsC 2 |
|  |  | Yes, by family doctor | RAblsC 3 |
|  |  | Yes, by professional first responder | RAblsC 4 |
|  |  | Yes, by civil first responder | RAblsC 5 |
|  |  | Yes, unknown by whom | RAblsC 7 |
| Initial Rhythm Shockable | Unknown | Unknown/missing | R1typeS 0 or N/A |
|  | No | No, supraventricular tachycardia | R1typeS 3 |
|  |  | No, normal rhythm | R1typeS 4 |
|  |  | No, bradycardia | R1typeS 5 |
|  |  | No, asystole | R1typeS 6 |
|  |  | No, unknown | R1typeS 7 |
|  | Yes | Yes, ventricular fibrillation | R1typeS 1 |
|  |  | Yes, ventricular tachycardia | R1typeS 2 |

##### Confidence intervals instead of *p*-values

Given that so-called ‘nominal *p*-values’ that are sometimes reported cannot be interpreted as *p*-values (as their distribution under the null sampling process is not (sub)uniform) we prefer to report no *p*-values. Our preferred inference is based on the corrected 95% confidence intervals accompanied by extensive descriptives of uncorrected/raw data results in tables.

##### Sample size re-estimation

Part of the sample size plan is to recalculate the maximum sample size of 2,400 patients at the 6th interim analysis after inclusion of 2,000 patients to decide whether a new trial should be prepared to increase the sample size beyond this trial. This sample size re-estimation will be performed based on blinded group allocation, but with knowledge of the two possible efficacy analyses (coded as A/B, B/A). In case a new trial starts, it will be analyzed in a prospective meta-analysis with this trial.

#### A.1.6 Estimands and role of supplementary analyses

Subsetting on observed intercurrent events in an mITT analysis does not, in general, capture a principal stratum estimand, but it does for our two intercurrent events of failure to initiate the intervention, no advanced airway and sustained mechanical ventilation. The estimate is unbiased because two conditions are met: first, the allocation does not influence the intercurrent events of airway placement and mechanical ventilation (no ‘intervention initiators’ or ‘control initiators’). Second, the intercurrent events of advanced airway placement and mechanical ventilation are clearly defined and measurable [41]. These are plausible: first decisions regarding advanced airway placement and mechanical ventilation are determined by EMS protocols, the provider’s airway experience, and patient need, rather than treatment allocation, which is blinded. In other words, being randomized to the PEEP or sham arm is highly unlikely to influence whether a patient receives advanced airway management or mechanical ventilation. Second, these decisions are consistently documented in ambulance records, ensuring transparency and reliable data.

Note that these conditions do not necessarily hold for the intercurrent event of study valve removal, which can be related to perceived valve results and is more difficult to measure. The subset of subjects who had their valve removed if allocated to PEEP could be different from those allocated to sham. As clearly highlighted in ICH E9(R1) addendum on estimands [42, p.10], including such an intercurrent event in our definition of the mITT subset is not recommended because it can confound the effects of the valve with the differences in outcomes possibly due to the differing characteristics of the patients.

**Figure A.4.**
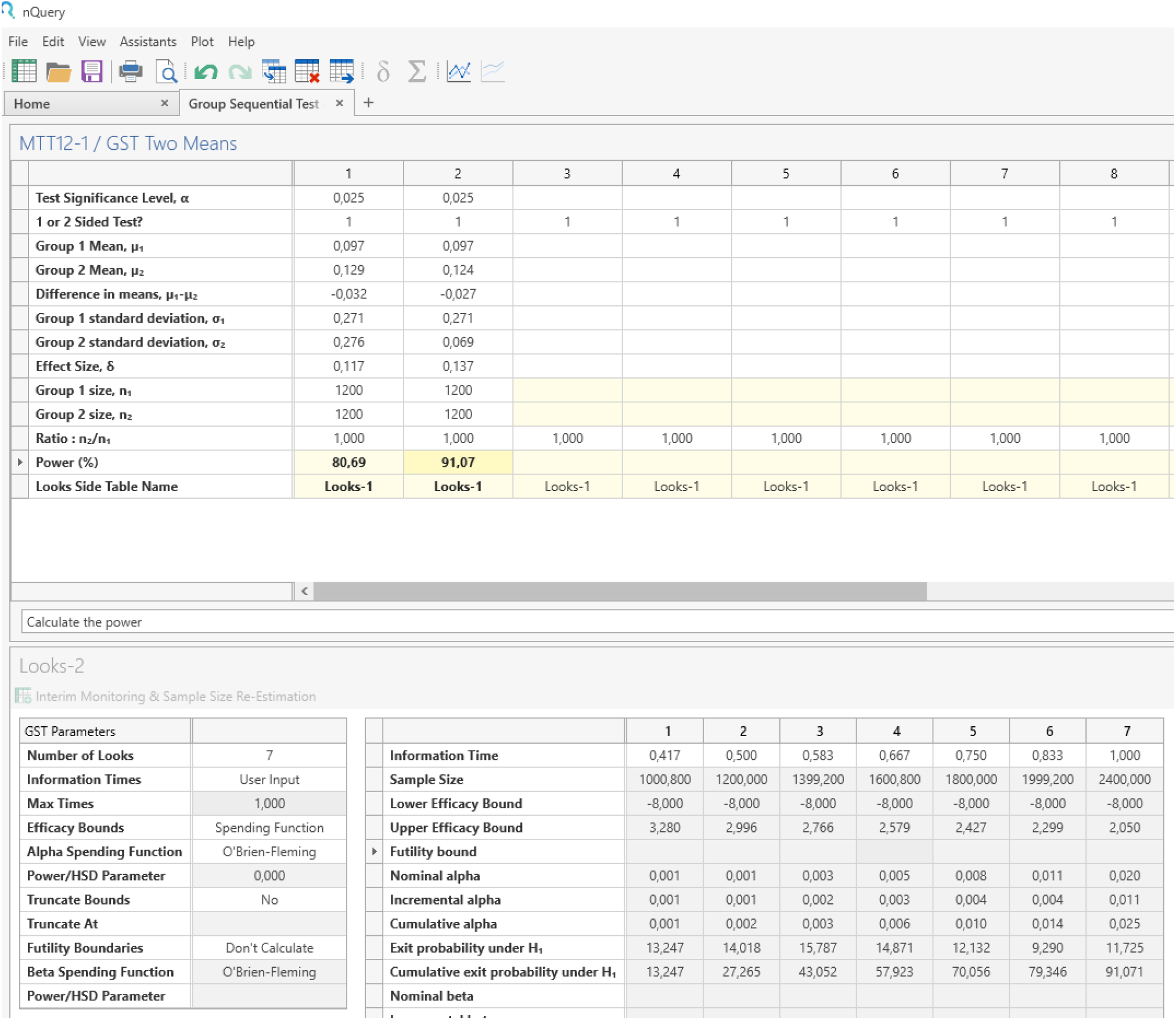
Power results from nQuery for a Cohen’s d of 0.1172 (Unfavorable scenario) and 0.1369 (Favorable scenario), with the Cohen’s d 0.1369 Favorable scenario illustrated in detail in the Looks-2 tables at the bottom.

##### Supplementary analyses

We do not consider the intention-to-treat (ITT) and per-protocol (PP) analyses as sensitivity analyses because they target a different estimand (ITT) than our primary analysis on the mITT subset or cannot be 1:1 related to any estimand (PP): administering ZEEP by removing the valve after start of resuscitation may be related to poor tolerance of PEEP, making it unclear what the target population of the PP analysis is. We report the ITT and PP analysis particularly for the completeness readers are accustomed to and use the term ‘supplementary analyses’ suggested in the ICH E9(R1) addendum [42] for analyses on these non-primary estimands.

##### Primary mITT analysis

The mITT and not the ITT is the primary analysis because only those patients are guaranteed to receive PEEP with the constant airway sealing needed for pressure build-up. This makes the mITT analysis more powerful than the ITT analysis in which patients without SAD or ETT dilute the effect. The outcomes will only apply to patients with advanced airway support, but a benefit conclusion on the primary analysis will inform the recommendation of 8 cm H_2_O PEEP for the entire resuscitation in practice where the same bag is used for bag-valve-mask as well as SAD/ETT-bag. Despite this practical aspect, we consider a benefit conclusion in the mITT analysis regardless of the result in the ITT analysis. We a-priori specified an effect in the non-mITT subset to be unconvincing due to the a-priori low probability that PEEP shows an effect with bag-valve-mask ventilation (mechanistically unrealistic) and limited power in this small subset that combined increase the risk of a false positive.

#### A.1.7 Power analysis scenarios

We defined an absolute risk difference of 4% in mortality as minimally clinically important (see PCI-RR design planner table Section A.1.9) and considered two scenarios for the neurological outcomes of these additional 4% survivors, a Favorable scenario and an Unfavorable scenario. Both scenarios are explained graphically in Figure A.5.

**The Favorable scenario** is called a neuroprotective effect in [43] and in our setting would mean that 4% of the total patients that would have died in the ZEEP/sham valve group now survive with a neurological outcome of mRS-5, while the same number of patients that would have survived with neurological outcome mRS-5, now survive with mRS-4, and the same thing up to mRS-1. Given the very small percentage of patients that is known from the ARREST database to survive with mRS-0, we consider it unlikely that such an additional number (4% of the total) would survive with mRS-0, so we leave these at mRS-1. This pattern can be seen as a shift, although shifts are assumed elsewhere to either refer to a shift of median score in a rank model [43], or shift of a mean of a latent continuous variable in a proportional odds model [31].

**The Unfavorable scenario** spreads the additional 4% survivors that would have died in the ZEEP/sham valve group according to the survivor base rate observed in the ARREST database. Given that a part of those survive as mRS-5 with a utility score of 0 and the utility scores of mRS-4 to mRS-2 are more different from mRS-1 than mRS-0, the mean difference in utility is smaller in this Unfavorable scenario than in the Favorable scenario.

##### Scenario details

An Excel sheet is provided as supplementary material [6] to further illustrate how a Cohen’s d is calculated from these scenarios, and to allow users to define other scenarios. Note that we define the base rate mortality (mRS-6) as 88% within modified intention-to-treat (mITT) subset. Given that this concerns the subset of OHCA patients for which manual ventilation with advanced airway management (intubation or supraglottic airway device) is initiated, we increased the base rate mortality estimate from the ARREST data base (due to resuscitation time bias, as advanced airway management typically occurs during prolonged CPR) and scaled the survivor’s base rates accordingly.

#### A.1.8 Data exclusion

The flow diagram of patients in Figure 1 illustrates the two possible reasons for exclusion of a patient from the primary analysis. The first occurs at the stage of allocation, in case a patient underwent randomization in error and should be excluded based on the exclusion criteria above (*<*18 years of age, suspected traumatic arrest or suspected drowning). The second reason occurs at the stage of follow-up and relates to missing data on the variables used for the primary analysis: the score on the modified Rankin Scale at discharge and the covariates. Table A.4 shows that unknown or missing values are prespecified as part of the yes/no coding of the covariates and therefore are no reason for patient exclusion. So for the primary analysis, only a missing value on the modified Rankin Scale was relevant, which can happen if a patient was brought to a hospital that has no collaboration with the ARREST registry. In case the primary outcome was missing, a patient was excluded from the primary analysis.

#### A.1.9 PCI-RR design planner table

The philosophy of the trial was in a sense that of Salim Yusuf, Rory Collins and Richard Peto [44] of large and simple trials. The REVIVE-PEEP trial has only one central question and analysis, on the primary outcome, and the main analysis is symmetric in terms of benefit and harm, so we use the same type-I error rate of 2.5% for each side of the null hypothesis (see also the DSMB charter [4] for instructions for safety monitoring in terms of the primary outcome). So we present the Design Planner Table here in transposed form for this single question. For the sampling plan we refer to Section 2.6 (Sample size) and A.1.7 (Power analysis scenarios) and for the analysis plan to sections 2.8 (Statistical analysis), A.1.4 (Main effect covariates for the primary analysis), A.1.5 (Interim analyses), A.1.6 (Estimands and role of supplementary analyses), A.1.8 (Data exclusion), Section 3 (Results) and A.2 (Results details).

##### Theoretical question

Does applying 8 cm H_2_O PEEP during manual ventilation after placement of a supraglottic airway or endotracheal tube improve neurological outcomes by enhancing oxygenation and preventing atelectasis, or could it worsen outcomes by reducing venous return and cardiac output?

##### Patient-centered question

Does PEEP during manual ventilation in addition to the standard advanced life support protocol improve modified Rankin Scale (mRS) outcomes weighted by Chaisinanunkul et al. (2025) [24] utility weights?

Here, we report the ordinal modified Rankin Scale that defines a score of 5 as better than 6 (death) but amend it in the analysis such that it cannot conclude benefit in case a decrease in mRS-6 was accompanied by an increase in mRS-5 both weighted as having 0 utility. So our main analysis answers the patient-centered question by taking into account utility weights. The underlying mechanism of enhanced oxygenation could improve neurological outcomes only from mRS-6 to mRS-5 and have no utility for patients, in which case our analysis can never successfully conclude for benefit. Such a result will be shared and inform research on the mechanism but will be secondary to the negative patient-centered conclusion. Hence it will not make us recommend further studies or analyze our data differently.

##### Rationale for deciding the sensitivity of the test for confirming or disconfirming the hypothesis

The study defines an absolute reduction of 4% of patients with mRS-6 (death) at discharge as the minimally clinically important difference between the control and intervention groups, based on the rationale that an intra-cardiac arrest intervention should lead to a significant improvement in survival in order to justify a change in clinical practice guidelines [45]. However, the cost and burden of the implementation of PEEP valves in manual ventilation are low, while the gain of each percentage point benefit is high, equaling hundreds of lives saved in the Netherlands and tens of thousands worldwide.

So we will recommend to implement 8 cm H_2_O PEEP also in case of an effect smaller than 4% as long as the data is only compatible with positive values for the difference in mean utility in terms of our pre-specified definition of a 95%-confidence interval. If we find no safety concerns on the occurrence of clinically significant pneumothorax, and an inconclusive result on the mean utility difference (with effect of benefit and harm both in the confidence interval), we recommend further studies to draw conclusions in a prospective meta-analysis, while taking into account feasibility (e.g. value of information).

##### Interpretation given different outcomes

Positive values for the corrected mean difference in utility refer to patient-centered benefit, and point to enhanced oxygenation. Negative values refer to harm, and point to adverse cardiac effects on defibrillation efficiency.

##### Theory that could be shown wrong by outcomes

The theoretical question supports the design of the study, but cannot be answered by the study. The emergency care situation of OHCA does not allow for standardized, detailed measurements of oxygenation or cardiac output. If better neurological outcomes are observed in the PEEP group than in the ZEEP/sham group (conclusive benefit with a confidence interval containing only positive values), improved oxygenation is the most likely cause but cannot be confirmed. If worse neurological outcomes are observed in the PEEP group than in the sham group (conclusive harm with a confidence interval containing only negative values) improved oxygenation cannot be ruled out, but apparently overshadowed by other causes – most likely decreased cardiac output.

### A.2 Results details

The confidence interval follows from the prespecified 2.050 times the standard error of 0.0101, specified at the final sample size 2,400, with the standard error obtained from the regression model at this final analysis corrected for the prespecified covariates (see Table A.5 and Table A.4). Note that this beta estimate is different from the combined mean utility difference reported based on the uncorrected data in Table 2 (one is positive, one is negative). Both estimates are relatively close to 0 (see Figure 3 for scale) compared to the scenarios used for sample size estimation. Both the regression-corrected and the uncorrected analyses are inconclusive, as the uncorrected standard error (0.0114) is larger than 0.0101 from the prespecified regression model, see Table A.5. Figure 3 also illustrates the six interim analyses that preceded this final analysis.

**Table A.5.** Neurological outcomes on the modified Rankin Scale weighted by Table 1: regression model with covariates from Appendix Table A.4 at the final analysis of 2,400 patients with mock results.

| Variable | Estimate | Std. Error |
| --- | --- | --- |
| (Intercept) | -0.0729340866 | 0.09326794 |
| interventionPEEP | 0.0072518211 | 0.01009323 |
| witnessOfCA_no-7 | -0.0002565743 | 0.08420449 |
| witnessOfCA_yesEMS-1 | 0.1828605893 | 0.08681498 |
| witnessOfCA_yesNonEMS-234568 | 0.0402213692 | 0.08404760 |
| bystanderCPR_no-6 | 0.0193672519 | 0.05516523 |
| bystanderCPR_yes-123457 | 0.0399881413 | 0.05382775 |
| initialRhythmshockable_no-34567 | 0.0172347625 | 0.05435212 |
| initialRhythmshockable_yes-12 | 0.2938520215 | 0.05464232 |

\*By adding in a linear model the covariates 'Witness of arrest', 'Bystander CPR' and 'Initial rhythm' as main effects with definitions as preregistered in the Stage-1 Registered Report, for the Favorable scenario, the pooled SD is expected to be reduced to 0.266–0.278 and Cohen's d increased to 0.131–0.137 and for the Unfavorable scenario, the pooled SD is expected to be reduced to 0.265–0.274 and Cohen's d increased to 0.117–0.121. SD = standard deviation

**Figure A.5.**
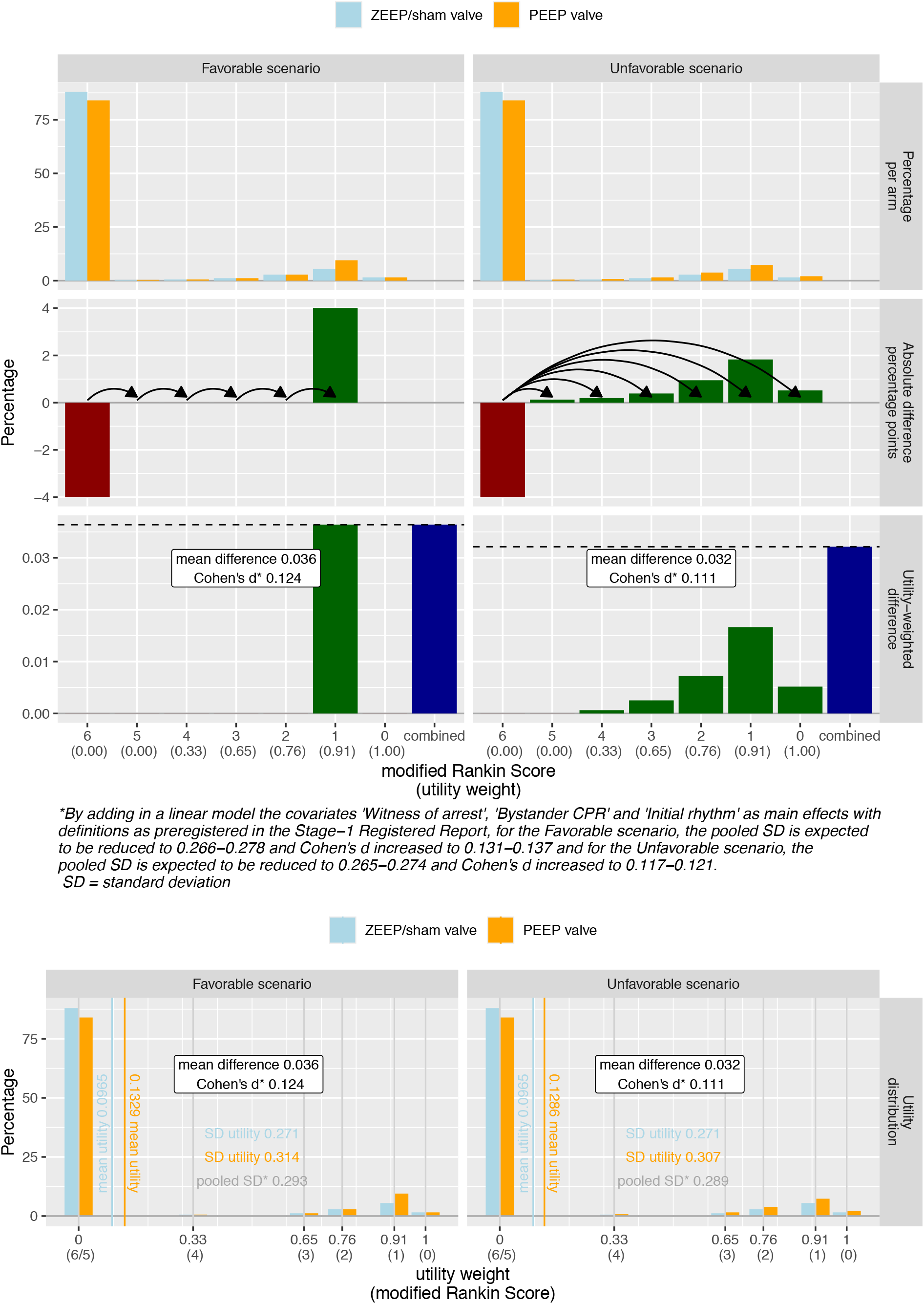
Definitions of the Favorable and Unfavorable scenario for neurological outcomes given the same 4%-point absolute decrease in mortality (mRS-6) and base rates of the survivor’s scores (mRS-0 to mRS-5) observed in the ARREST registry period 2016-2018. These base rates might overestimate the proportion of good outcomes as withholding consent for mRS-scores to be included in research was found to be associated with worse outcomes elsewhere [46]. Figure based on Excel sheet [6] using R code [7].

## Footnotes

1 The ethics committee that received the submission for the REVIVE-PEEP trial on February 13, 2026 (scheduled for their meeting on February 26) was METC Amsterdam UMC. This ethics committee merged with the METC NedMec on July 1, therefore the ethics approval on July 22 was granted by METC NedMec. The new name of the merged committees is ‘METC NedMec+’ but the public registry shows ethics approval by ‘METC NedMec’. Information on the merger is available in English: https://toetsingonderzoek.amsterdamumc.org/

2 The review process at the Dutch Heart Foundation took place in April 2026 and the awarded grant was press-released on July 23, 2026: https://professionals.hartstichting.nl/actualiteiten/klinische-studie-naar-beademing-bij-reanimaties-buiten-het-ziekenhuis

3 https://onderzoekmetmensen.nl/nl/trial/61400

## References

[1] Judith ter Schure. Replication Package REVIVE-PEEP trial Registered Report. ResearchEquals, 2026. 10.53962/p8t5-wzwr.

[2] Jeroen van Eijk, Judith ter Schure, Michiel Hulleman, Stephan A. Loer, Lothar A. Schwarte, Thijs Delnoij, Hans van Schuppen, and Patrick Schober. REVIVE-PEEP trial research protocol. medRxiv, 2026. 10.64898/2026.02.26.26346617.

[3] Jeroen van Eijk. REVIVE-PEEP - Participant Information Sheet Version 2.2. ResearchEquals, 2026. 10.53962/t6cr-dxh3.

[4] Judith ter Schure and Jeroen van Eijk. REVIVE-PEEP Protocol Stage-1 Registered Report Supplement charter Data and Safety Monitoring Board (DSMB) Version 2.1. ResearchEquals, 2026. 10.53962/zdxb-p0m2.

[5] Judith ter Schure, Mette Ekkel, and Remy Stieglis. REVIVE-PEEP Protocol Stage-1 Registered Report Supplement example DSMB closed session report with mock data. ResearchEquals, 2026. 10.53962/vcy9-0xar.

[6] Judith ter Schure. REVIVE-PEEP Protocol Stage-1 Registered Report Supplement Excel sheet to inspect various scenarios (modified Rankin Scale → Cohen’s d) based on Chaisinanunkul et al. (2015) utility weights. ResearchEquals, 2026. 10.53962/pk22-sgne.

[7] Judith ter Schure. REVIVE-PEEP Protocol Stage-1 Registered Report Supplement R code to create scenario plot based on Excel sheet. ResearchEquals, 2026. 10.53962/t3sn-vqqr.

[8] Yuki Mori, Kaito Takashima, Kohei Ueda, Kyoshiro Sasaki, and Yuki Yamada. Trinity review: integrating registered reports with research ethics and funding reviews. BMC research notes, 15(1):184, 2022.

[9] MT Blom, DA Van Hoeijen, A Bardai, J Berdowski, PC Souverein, ML De Bruin, RW Koster, A De Boer, and Hanno L Tan. Genetic, clinical and pharmacological determinants of out-of-hospital cardiac arrest: rationale and outline of the AmsteRdam RESsuscitation STudies (ARREST) registry. Open heart, 1(1), 2014.

[10] Jan-Thorsten Gräsner, Jan Wnent, Johan Herlitz, Gavin D Perkins, Rolf Lefering, Ingvild Tjelmeland, Rudolph W Koster, Siobhán Masterson, Fernando Rossell-Ortiz, Holger Maurer, et al. Survival after out-of-hospital cardiac arrest in europe-results of the eureca two study. Resuscitation, 148:218–226, 2020.

[11] Jeroen A van Eijk, Lotte C Doeleman, Stephan A Loer, Rudolph W Koster, Hans van Schuppen, and Patrick Schober. Ventilation during cardiopulmonary resuscitation: A narrative review. Resuscitation, 203:110366, 2024.

[12] Walter Spindelboeck, Otmar Schindler, Adrian Moser, Florian Hausler, Simon Wallner, Christa Strasser, Josef Haas, Geza Gemes, and Gerhard Prause. Increasing arterial oxygen partial pressure during cardiopulmonary resuscitation is associated with improved rates of hospital admission. Resuscitation, 84(6):770–775, 2013.

[13] Walter Spindelboeck, Geza Gemes, Christa Strasser, Kathrin Toescher, Barbara Kores, Philipp Metnitz, Josef Haas, and Gerhard Prause. Arterial blood gases during and their dynamic changes after cardiopulmonary resuscitation: a prospective clinical study. Resuscitation, 106:24–29, 2016.

[14] Jignesh K Patel, Elinor Schoenfeld, Puja B Parikh, and Sam Parnia. Association of arterial oxygen tension during in-hospital cardiac arrest with return of spontaneous circulation and survival. Journal of intensive care medicine, 33(7):407–414, 2018.

[15] Jasmeet Soar, Bernd W Böttiger, Pierre Carli, Francesc Carmona Jiménez, Diana Cimpoesu, Gareth Cole, Keith Couper, Sonia D’Arrigo, Charles D Deakin, Jacqueline Eleonora Ek, et al. European resuscitation council guidelines 2025 adult advanced life support. Resuscitation, 215:110769, 2025.

[16] NJ Johnson, G Debaty, BY Yang, A Moskowitz, I Drennan, J del Castillo, JE Bray, T Olasveengen, LJ Morrison, on behalf of the International Liaison Committee on Resuscitation Basic Life Support, and Advanced Life Support Task Forces. Ventilation parameters during adult cardiopulmonary resuscitation: Consensus on science with treatment recommendations. International Liaison Committee on Resuscitation (ILCOR), 2026.

[17] Tom P Aufderheide, Graham Nichol, Thomas D Rea, Siobhan P Brown, Brian G Leroux, Paul E Pepe, Peter J Kudenchuk, Jim Christenson, Mohamud R Daya, Paul Dorian, et al. A trial of an impedance threshold device in out-of-hospital cardiac arrest. The New England journal of medicine, 365, 2011.

[18] Yosef Levenbrown, Md Jobayer Hossain, James P Keith, Katlyn Burr, Anne Hesek, and Thomas Shaffer. The effect of positive end-expiratory pressure on cardiac output and oxygen delivery during cardiopulmonary resuscitation. Intensive Care Medicine Experimental, 8(1):36, 2020.

[19] Miriam Renz, Leah Müllejans, Julian Riedel, Katja Mohnke, René Rissel, Alexander Ziebart, Bastian Duenges, Erik Kristoffer Hartmann, and Robert Ruemmler. High peep levels during cpr improve ventilation without deleterious haemodynamic effects in pigs. Journal of Clinical Medicine, 11(16):4921, 2022.

[20] Jukka Kopra, Erik Litonius, Pirkka T Pekkarinen, Merja Laitinen, Juho A Heinonen, Luca Fontanelli, and Markus B Skrifvars. Oxygenation and ventilation during prolonged experimental cardiopulmonary resuscitation with either continuous or 30: 2 compression-to-ventilation ratios together with 10 cmh20 positive end-expiratory pressure. Intensive care medicine experimental, 12(1):36, 2024.

[21] Ricardo Luiz Cordioli, Laurent Brochard, Laurent Suppan, Aissam Lyazidi, Francois Templier, Abdo Khoury, Stephane Delisle, Dominique Savary, and Jean-Christophe Richard. How ventilation is delivered during cardiopulmonary resuscitation: an international survey. Respiratory care, 63(10):1293–1301, 2018.

[22] Kirsty Loudon, Shaun Treweek, Frank Sullivan, Peter Donnan, Kevin E Thorpe, and Merrick Zwarenstein. The PRECIS-2 tool: designing trials that are fit for purpose. BMJ, 350, 2015.

[23] Jeroen van Eijk, Patrick Schober, Hans van Schuppen, and Judith ter Schure. Positive end-expiratory pressure versus sham valve/zero end-expiratory pressure in cardiopulmonary resuscitation during manual ventilation to improve neurological outcomes in adult patients suffering an out-of-hospital cardiac arrest – an investigator-initiated, pragmatic, registry-based, multicenter, parallel-group, tripleblind randomized controlled superiority clinical trial in the ARREST registry (REVIVE-PEEP protocol Stage-1 Registered Report). medRxiv, 2026. 10.64898/2026.08.27.26361533.

[24] Napasri Chaisinanunkul, Opeolu Adeoye, Roger J Lewis, James C Grotta, Joseph Broderick, Tudor G Jovin, Raul G Nogueira, Jordan J Elm, Todd Graves, Scott Berry, et al. Adopting a patient-centered approach to primary outcome analysis of acute stroke trials using a utility-weighted modified Rankin scale. Stroke, 46(8):2238–2243, 2015.

[25] Jan-Thorsten Grasner, Janet E Bray, Jerry P Nolan, Taku Iwami, Marcus E H Ong, Judith Finn, Bryan McNally, Ziad Nehme, Comilla Sasson, Janice Tijssen, Shir Lynn Lim, Ingvild Tjelmeland, Jan Wnent, Bridget Dicker, Chika Nishiyama, Zakary Doherty, Michelle Welsford, Gavin D Perkins, and International Liaison Committee on Resuscitation. Cardiac arrest and cardiopulmonary resuscitation outcome reports: 2024 update of the Utstein Out-of-Hospital Cardiac Arrest Registry template. Resuscitation, 201:110288, 2024.

[26] Céline Occelli, Marie Lenoir, Arthur Naudet Lasserre, Lauranne Teule, Hugues Weber, Jonathan Charbit, and Xavier Bobbia. Prehospital diagnostic performance of emergency physicians in identifying blunt traumatic pneumothorax requiring early decompression. BMC Emergency Medicine, 26(1):49, 2026.

[27] Lianne JP Sonnemans, Alireza R Bayat, Aniek RC Bruinen, Marleen H Van Wely, Marc A Brouwer, Dennis Bosboom, Johannes G Van Der Hoeven, Mathias Prokop, and Willemijn M Klein. Comparing thoracoabdominal injuries of manual versus load-distributing band cardiopulmonary resuscitation. European Journal of Emergency Medicine, 27(3):197–201, 2020.

[28] Berry Consultants and Scott Berry. “In the Interim… ” podcast Episode 5: Religion, Politics, and Ordinal Outcomes. https://www.berryconsultants.com/resource/5-religion-politics-and-ordinal-outcomes, March 24, 2025.

[29] Berry Consultants, Scott Berry, and Jeffry Saver. “In the Interim… ” podcast Episode 37: A Visit with Stroke Neurologist Dr. Jeff Saver. https://www.berryconsultants.com/resource/37-a-visit-with-stroke-neurologist-dr-jeff-saver, November 17, 2025.

[30] Berry Consultants, Scott Berry, and Linsay Berry. “In the Interim… ” podcast Episode 50: The Fallacy of Ordinal Endpoints. https://www.berryconsultants.com/resource/50-the-fallacy-of-ordinal-endpoints, February 23, 2026.

[31] Yongxi Long, Eveline JA Wiegers, Bart C Jacobs, Ewout W Steyerberg, and Erik W van Zwet. Role of the proportional odds assumption for the analysis of ordinal outcomes in neurologic trials. Neurology, 105(8):e214146, 2025.

[32] Posit team. RStudio: Integrated Development Environment for R. Posit Software, PBC, Boston, MA, 2026.

[33] Hadley Wickham. ggplot2: Elegant Graphics for Data Analysis. Springer-Verlag New York, 2016.

[34] Hadley Wickham, Romain François, Lionel Henry, Kirill Müller, and Davis Vaughan. dplyr: A Grammar of Data Manipulation, 2023. R package version 1.1.4.

[35] Baptiste Auguie. gridExtra: Miscellaneous Functions for “Grid” Graphics, 2017. R package version 2.3.

[36] Mathias Legrand. Stylish Article LaTeX Template Version 2.0, 2014. CC BY-NC-SA 3.0, (13/4/14); Accessed via Overleaf in 2025/2026: https://www.overleaf.com/latex/templates/stylish-article-template/grgwqvchdmns.

[37] Arthur Bouillon, Maxim Vanwulpen, Thomas Tackaert, Ruben Cornelis, and Said Hachimi-Idrissi. Explorative study on lower inflection point dynamics during cardiopulmonary resuscitation: Potential implications for airway management. Resuscitation, 200:110242, 2024.

[38] Mathini Vaseekaran, Tobias Vollmer, Lydia Johnson Kolaparambil Varghese, Christina Zeichen, Jochen Hinkelbein, Raphael Abels, Bernd Strickmann, Martin Deicke, Julia Johanna Grannemann, Rainer Grünzig, et al. Peri-arrest ventilation with positive endexpiratory-pressure vs. zero end-expiratory-pressure in out-of-hospital-cardiac-arrest (peravent)—a prospective, cluster-randomized multicenter trial. Trials, 27(1):69, 2026.

[39] Gerrit Jansen and Jochen Hinkelbein. PEEP vs. ZEEP in Out-of-Hospital-Cardiac-Arrest (PerAVent). https://clinicaltrials.gov/study/NCT06836830?term=NCT06836830&rank=1, 2025. ClinicalTrials.gov Identifier: NCT06836830. Updated August, 6, 2025. Accessed February 25, 2026.

[40] Thomas Tackaert. The Application of Positive End-Expiratory Pressure in Out-of-Hospital Cardiac Arrest: The Lazarus-PEEP Trial. (Lazarus-PEEP). https://clinicaltrials.gov/study/NCT06939335?term=NCT06939335&rank=1, 2025. ClinicalTrials.gov identifier: NCT06939335. Updated January, 21, 2026. Accessed February 25, 2026.

[41] Brennan C Kahan, Ian R White, Mark Edwards, and Michael O Harhay. Using modified intention-to-treat as a principal stratum estimator for failure to initiate treatment. Clinical Trials, 20(3):269–275, 2023.

[42] Committee for Medicinal Products for Human Use et al. European medicines agency. ich e9 (r1) addendum on estimands and sensitivity analysis in clinical trials to the guideline on statistical principles for clinical trials. amsterdam, the netherlands: European medicines agency; 2020, 2023.

[43] Jeffrey L Saver and Jeffrey Gornbein. Treatment effects for which shift or binary analyses are advantageous in acute stroke trials. Neurology, 72(15):1310–1315, 2009.

[44] Salim Yusuf, Rory Collins, and Richard Peto. Why do we need some large, simple randomized trials? Statistics in medicine, 3(4):409–420, 1984.

[45] Graham Nichol, Siobhan P Brown, Gavin D Perkins, Francis Kim, Fritz Sterz, Jo Ann Broeckel Elrod, Spyros Mentzelopoulos, Richard Lyon, Yaseen Arabi, Maaret Castren, et al. What change in outcomes after cardiac arrest is necessary to change practice? results of an international survey. Resuscitation, 107:115–120, 2016.

[46] Joshua G Salzman, Ralph J Frascone, Nathan Burkhart, Richard Holcomb, Sandi S Wewerka, Robert A Swor, Brian D Mahoney, Marvin A Wayne, Robert M Domeier, Michael L Olinger, et al. The association of health status and providing consent to continued participation in an out-of-hospital cardiac arrest trial performed under exception from informed consent. Academic Emergency Medicine, 22(3):347–353, 2015.

